# Impact and cost-effectiveness of frequent HIV re-testing among key populations in Viet Nam: a modeling study

**DOI:** 10.1101/2022.03.01.22271733

**Authors:** Dylan Green, David Coomes, Ruanne V. Barnabas, Monisha Sharma, Magdalena Barr-Dichiara, Muhammad S. Jamil, Morkor Newman Owiredu, Virginia Macdonald, Van Nguyen, Son Vo Hai, Theodora E. Wi, Cheryl Johnson, Alison L. Drake

## Abstract

**Background:** HIV testing and counseling is a key component of HIV prevention and the entry point into the treatment cascade, which improves for individual clinical outcomes and reduces onward HIV transmission. Current guidelines recommend at least annual testing for key populations. More frequent testing could provide health benefits, but these additional services increase the program the cost-effectiveness is not well-evaluated.

**Methods:** We used a compartmental mathematical model to simulate the health and economic impact of HIV testing one to four times per year for men who have sex with men, people who inject drugs, and female sex workers in Viet Nam. Model outcomes included costs, HIV infections, HIV-related deaths, and disability-adjusted life years (DALYs) associated with each scenario. We used an opportunity cost-based cost-effectiveness threshold of US $2,255 per DALY averted, discounted costs and health benefits at 3% annually, and used a time horizon from 2021 to 2030 to calculate incremental cost-effectiveness ratios (ICERs).

**Results:** Compared to the baseline scenario, more frequent HIV testing was estimated to incrementally avert 10.2%, 5.2%, 3.0%, and 1.6% discounted infections for one-, two-, three-, and four-tests per year, respectively. ICERs associated with each scenario ranged from $464, $1,190, $1,762, and $2,727 per DALY averted for one-, two-, three-, and four-tests per year, respectively.

**Conclusions:** Increased HIV testing frequency for key populations was projected to avert HIV incidence, mortality, and disability in Viet Nam and was cost-effective. Settings with a similar context should consider strategies on how to optimize retesting among key populations.

## Background

The World Health Organization (WHO) recommends initiation of antiretroviral treatment (ART) for all persons testing positive for HIV regardless of CD4 count and the United Nations Joint Programme on HIV/AIDS (UNAIDS) put forth the 95-95-95 goals which seek to ensure 95% of persons living with HIV are aware of their status, provide ART for 95% of those diagnosed, and maintain viral suppression among 95% persons living with HIV on ART by 2030.^1,2^ Although mathematical models of the 95-95-95 goals predict large declines in HIV incidence, empirical estimates of HIV incidence reductions have been less than expected, and that feasibility and costs present challenges in attaining such ambitious targets.^3–7^ Late diagnosis and delayed ART initiation are key barriers to reaching HIV incidence reduction goals, as acute infections become a larger proportion of total transmission as ART is scaled up.^4,8^

Key populations including men who have sex with men, people who inject drugs, and female sex workers are disproportionately impacted by HIV, comprising over 60% of new HIV infections globally with a lower proportion tested, linked to care, and virally suppressed compared to the general population.^9–11^ While data on knowledge of HIV positive status among key populations are limited, UNAIDS estimates of that between 62-67% of key populations living with HIV globally are aware of their status compared to 81% for all persons living with HIV.^9,12^

Current WHO HIV testing guidelines recommend retesting at least annually for key populations.^13^ However, the optimal testing frequency for high incidence populations to rapidly diagnose and treat individuals is not well evaluated. More frequent HIV retesting strategies could increase the rate of persons living with HIV diagnosed and initiated on ART, and maybe required to reach the goal of 95% of persons living with HIV aware of their status.^14^ However, more frequent testing will also increase program costs through additional commodity procurement, as well as increasing health systems burden, program coordination, and outreach. There are also opportunity costs to consider, as resources allocated to more frequent HIV testing could be used for a competing health need.

Viet Nam has a national HIV prevalence of less than 1% in the general population, but prevalence ranges between 3-25% among men who have sex with men, people who inject drugs, and female sex workers.^15,16^ While Viet Nam has made universal ART with financing through its social health insurance scheme and the Global Fund to Fight AIDS, Tuberculosis and Malaria, ART coverage stands at 69%. This suboptimal ART coverage could be due to incomplete linkage to care among the newly diagnosed and loss to follow-up. Another key barrier to universal ART could be a low rate of diagnosis. Furthermore, the HIV funding landscape in Viet Nam has been transitioning from donor to national social health insurance which has a mandate to meet priority health needs including HIV.^17^ The cost-effectiveness and impact of health interventions provide evidence for policy makers to support efficient resource allocation. In this study, we modeled the health and economic impact of increased HIV testing frequency in key populations in Viet Nam to identify the most cost-effective frequency for retesting.

## Methods

### Setting and Populations

The target populations for this modeling analysis were adults aged 15 and greater who are people who inject drugs, female sex workers, and men who have sex with men in Viet Nam. HIV prevalence ranged from 12.7%, 3.6%, and 10.8% respectively (Table 1).

**Table 1.** Summary of key model parameters

| Input | Value | Sources and Notes |
| --- | --- | --- |
| **HIV Prevalence** |  |  |
| Men Who Have Sex with Men | 10.8% | HSS+2018<sup>15</sup> |
| People Who Inject Drugs | 12.7% | HSS+2019<sup>16</sup> |
| Female Sex Workers | 3.6% | HSS+2018<sup>15</sup> |
| **ART** |  |  |
| 2020 Coverage | 69% | AEM TWG |
| Annual Scale-Up | 3.0% | Strategic Plan Targets |
| Transmission Reduction Efficacy | 93% | HPTN 052<sup>34</sup> |
| Mortality Reduction Efficacy | Varies* | leDEA Consortium<sup>35</sup> |
| **Other Prevention** |  |  |
| Condom Efficacy | 80% | Cochrane Review<sup>36</sup> |
| Condom Use – Men Who Have Sex with Men | 50% | HSS+2018<sup>15</sup> |
| Condom Use – Female Sex Workers | 80% | HSS+2018<sup>15</sup> |
| 2020 PrEP Coverage – Men Who Have Sex with Men | 6% | Strategic Plan Targets |
| 2025 PrEP Coverage – Men Who Have Sex with Men | 23% | Strategic Plan Targets |
| PrEP Efficacy | 90% | PROUD and IPERGAY<sup>37,38</sup> |
| PrEP Adherence | 80% | Assumption |
| **Costs** |  |  |
| HIV Lay Test | \$4.50 | Expert Opinion |
| ART | \$285 | Expert Opinion |
| Time Horizon | 2020-30 | Strategic Plan Timeline |
| Discount Rate – Costs and Health Benefits | 3% | HTA guidelines. |

ART, antiretroviral therapy; PrEP, pre-exposure prophylaxis; HSS+, HIV sentinel surveillance; HPTN, HIV prevention trials network; AEM TWG, Asian Epidemic Model Technical Working Group; leDEA, International epidemiology Databases to Evaluate AIDS; HTA, Health Technology Assessment.
\* Mortality reduction due to ART varies by age, CD4 count, and time on ART

### Model

We used the AIDS Impact Model within the Spectrum software package (v 6.1) to simulate the HIV epidemic in Viet Nam.^18^ The model is a deterministic, compartmental mathematical model that simulates HIV transmission vertically from mother-to-child, intravenously via needle sharing, and hetero- and homosexual behavior and estimates HIV incidence, AIDS-related mortality, and disability-adjusted life years (DALYs). DALY estimates are stratified by adults and children, CD4 count (<200 vs ≥200 cells/µL) and ART use. The model is age- and sex-stratified with compartments for risk groups such as men who have sex with men, people who inject drugs, female sex workers and their clients, and low and medium risk heterosexuals.

The model was parameterized with demographic, behavioral, transmission, programmatic, epidemiological and efficacy data from government sources, surveys, surveillance, publicly available reports, databases, and peer-reviewed literature. The model was calibrated to risk group-specific HIV prevalence estimates for 2018-2019, total persons living with HIV, and HIV incidence within 1% of national estimates. We simulated the impact of increasing HIV retesting frequency using the Goals model within Spectrum.

### Scenarios

The current national guidelines for HIV testing in Viet Nam recommend testing every six months for key populations. However, current utilization is estimated at approximately 50% testing annually and therefore falls short of that guideline. We use this level as the baseline scenario until 2030 – the time horizon for Viet Nam’s national HIV strategic plan.^19^ We also simulated model scenarios with more frequent HIV retesting from one- to four-times per year among key populations from 2021 until 2030. HIV testing and retesting has the potential to impact the HIV epidemic in two ways: 1) increasing diagnosis and ART enrollment, and 2) resulting in behavior change including reduced number of sexual partners, increased condom use, and decreased needle sharing. Results of a systematic review of the effects of HIV testing on behavior yielded inconsistent findings, so we focus on the impact of HIV testing on ART enrollment.^20^ However, data on impact of HIV testing on ART uptake, especially on retesting, is extremely limited. Therefore, we made conservative assumptions on the linkage between testing frequency and ART use. Counterfactual to no increase in HIV testing, we assume that ART coverage meets the targets of the National Strategic Plan in 2025, and then continues to increase at historical averages until 95% coverage is met in 2030, approximately an average increase of 3% per year. We expect ART coverage to increase with scenarios of one-, two-, three-, and four-times per year. We estimate the largest impact of HIV testing on ART scale-up is to occur as one moves from never testing to a single test per year, with a decay in the marginal gains of additional annual tests (Figure 1). We assume that ART use reduces transmission risk by 70% and reduces mortality risk by 80% with an average 12-month retention of 83%.

**Figure 1:**
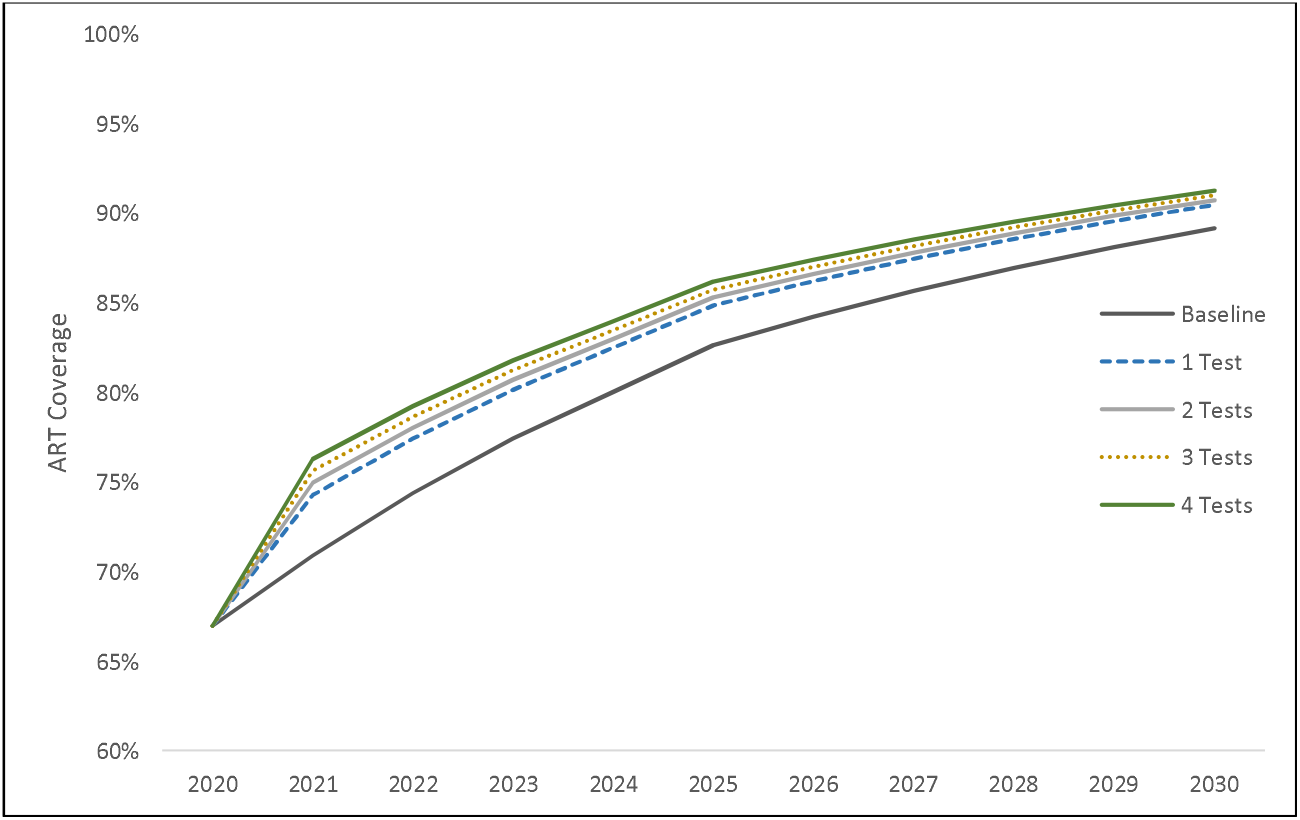
Assumed ART Scale-Up by HIV Test Policy Scenario

We also assume condoms are 80% efficacious and that use among men who have sex with men is 50% and 80% among female sex workers. We assume 55% of people who inject drugs access needle and syringe exchange. We assume PrEP efficacy is 90% and adherence is 80%, while scale-up and utilization is less than 2% among female sex workers and people who inject drugs. For men who have sex with men we assume PrEP coverage increases to 6% by 2020 and reaches 23% by 2025 (in alignment with targets) and is constant up to 2030.

### Costs

Costs were estimated from the provider perspective. We used local data on the personnel, commodities, and transport costs associated with testing and estimate the cost per HIV test at US $4.50. ART costs were obtained from local sources and included personnel, commodities, clinical follow-up, and laboratory monitoring for an annual cost of US $285, and we assumed individuals were on ART for an average of 25 years. For estimating net costs, the additional incurred costs due to faster ART scale-up as well as averted costs of ART through infections averted are also considered.

We do not assume that all individuals in target key populations get tested, nor do we assume that all available tests are utilized. We assume that 25% of key populations do not get tested annually. We assume that a decreasing proportion of the testing population utilizes additional tests with each scenario. Briefly, we assume 75% of persons would be tested once per year under a one-test per year policy, while 68% would be tested twice under a two-test per year policy, 61% tested three times, and only 55% fully utilizing all four tests under a four-Test annually scenario (Figure 2). This generates an average number of tests per person of 0.5, 0.8, 1.4, 2.0, and 2.6 for Baseline, one, two, three, and four test per year scenarios.

**Figure 2:**
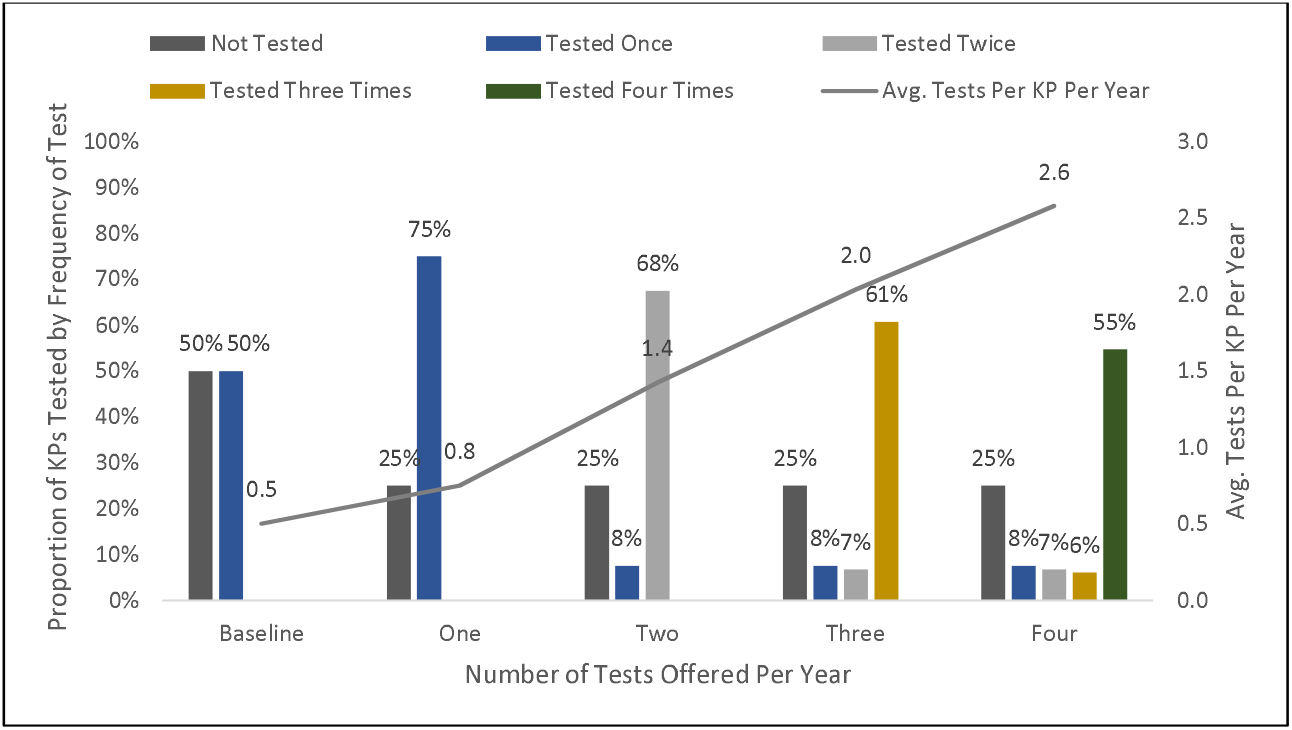
Testing Uptake by Test Frequency and Test Policy Scenario KP, key populations.

### Cost-Effectiveness Estimates, Thresholds, and Sensitivity Analysis

Health impact was measured in disability-adjusted life years (DALYs), infections, and AIDS-related deaths averted and aggregated over the 10-year time horizon. We use disability weights of 0.221, 0.547, and 0.053 for persons living with HIV with CD4 counts ≥200 cells/µL, <200 cells/µL, and on ART, respectively.^21^ Incremental costs were calculated as the costs of increasing testing frequency, ART costs incurred and averted because of testing. Costs and health benefits were discounted at 3% per year.

National Health Technology Assessment guidelines for Viet Nam are currently under development by local authorities. We used an opportunity-cost-based cost-effectiveness threshold, based on a method previously described.^22^ Briefly, the approach uses an empirical opportunity cost estimate along with a nation’s gross domestic product per capita and the value of a statistical life to refine the cost-effectiveness threshold. Using this approach, we considered a threshold of US $2,255 per DALY averted as cost-effective.

We conducted probabilistic sensitivity analyses on the following parameters: testing utilization, testing cost, ART cost, average years on ART, and intervention impact. We conducted 10,000 Monte Carlo simulations with random draws of parameter values across a specified distribution and report the proportion of simulated ICERs beyond our cost-effectiveness thresholds.

## Results

### Impact

Under the baseline scenario, we project a total of 43,886 new HIV infections and 14,503 AIDS-related deaths in Viet Nam from 2020 to 2030 (Table 2). We estimate that 3,953 (10.2%), 1,805 (5.2%), 1,002 (3.0%), and 498 (1.6%) new infections would be averted under policies of one-, two-, three-, and 4-tests per year, respectively (Figure 3). We estimate discounted AIDS-related deaths would be reduced by 616 (4.9%), 296 (2.5%), 166 (1.4%), and 92 (0.8%) for testing one-, two-, three-, and four-times per year, respectively. We further estimate that increased testing policies could avert 38,043, 18,124, 10,002, and 5,400 DALYs, respectively.

**Table 2:** Modeled Infections, Deaths, and DALYs Under Each Test Policy Scenario

|  | Baseline | 1 Test | 2 Tests | 3 Tests | 4 Tests |
| --- | --- | --- | --- | --- | --- |
| **Impact** |  |  |  |  |  |
| **Non-Discounted** |  |  |  |  |  |
| *New HIV* |  |  |  |  |  |
| *Infections* | 43,886 | 39,238 | 37,198 | 36,086 | 35,543 |
| *AIDS deaths* | 14,503 | 13,753 | 13,397 | 13,199 | 13,090 |
| **Discounted** |  |  |  |  |  |
| *Averted* |  |  |  |  |  |
| *New HIV* |  |  |  |  |  |
| *Infections* | - | 3,953<br>(10.2%) | 1,805 (5.2%) | 1,002 (3.0%) | 498 (1.6%) |
| *AIDS deaths* | - | 616 (4.9%) | 296 (2.5%) | 166 (1.4%) | 92 (0.8%) |
| *DALYs* | - | 38,043 | 18,124 | 10,002 | 5,400 |
| **Costs** |  |  |  |  |  |
| **Discounted** |  |  |  |  |  |
| Net Costs | \$12,305,781 | \$29,968,522 | \$40,007,289 | \$52,670,052 | \$64,704,300 |
| *Cost of HIV testing* | \$12,305,781 | \$18,440,443 | \$35,045,088 | \$49,981,897 | \$63,412,256 |
| *Cost of HIV* |  |  |  |  |  |
| *Treatment* | - | \$11,528,079 | \$4,962,201 | \$2,688,155 | \$1,292,044 |
| Incremental Cost | - | \$17,662,741 | \$21,566,846 | \$17,624,964 | \$14,722,403 |
| **ICER Per DALY Averted** | - | \$464 | \$1,190 | \$1,762 | \$2,727 |

DALY, disability-adjusted life year; ICER, incremental cost-effectiveness ratio.

**Figure 3:**
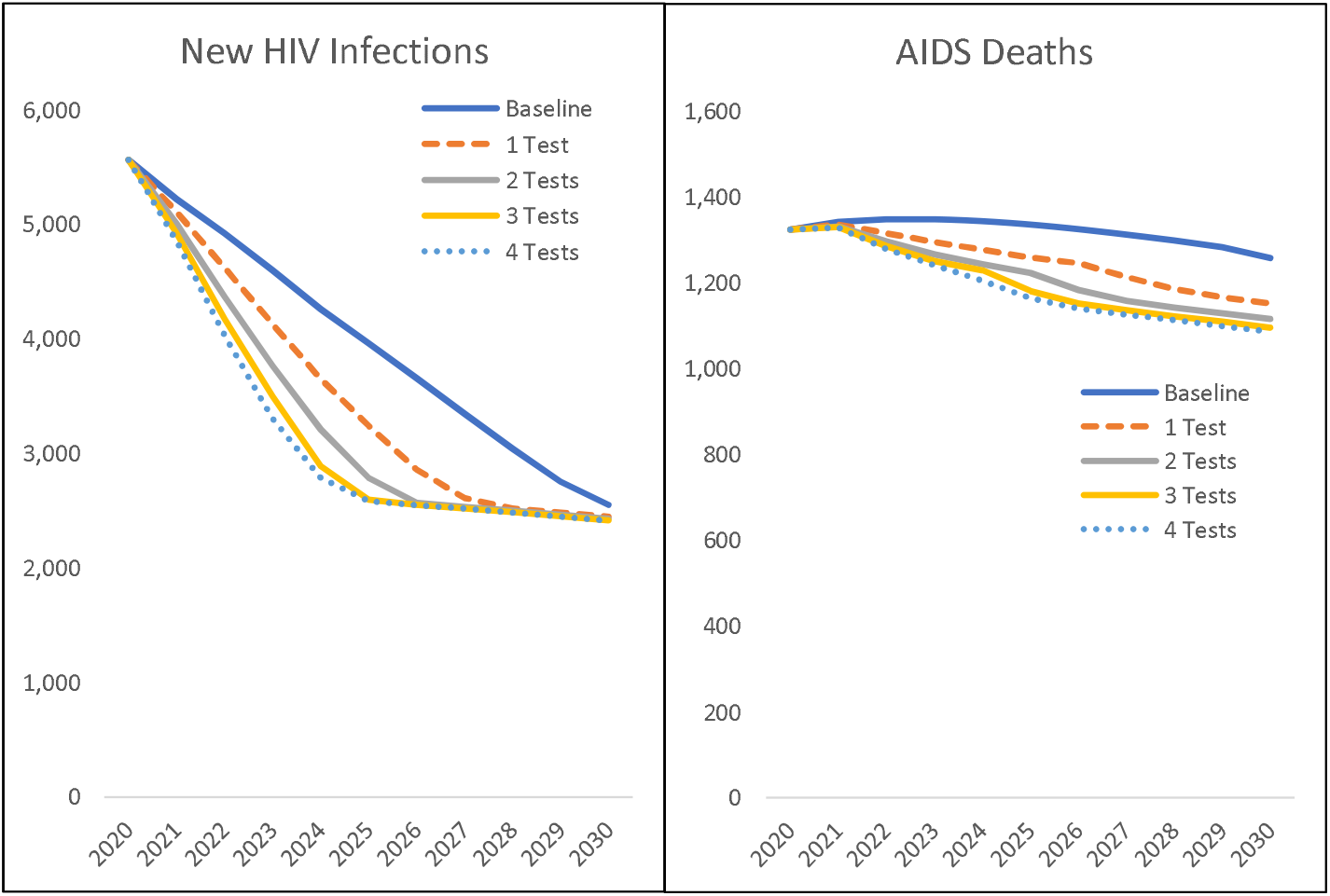
Modeled New HIV Infections and AIDS Deaths by Year and Test Policy Scenario

### Costs

At baseline, we estimate HIV testing costs approximately US $12.3 million. A one-test per year policy would cost an additional US $18.4 million, while incurring an additional US $11.5 million in net ART costs from 2020 to 2030. The cost of HIV testing increases almost linearly with each scenario, with some reduction due to less than complete utilization. We estimate testing costs to be US $35.0 million, US $49.9 million, and US $63.4 million for two-, three-, and four-tests per year scenarios, respectively. Conversely, net ART costs decline as treatment scale-up decelerates and additional costs are reduced through HIV infections averted, resulting in net ART costs of US $4.9 million, US $2.7 million, and US $1.3 million for two-, three-, and four-tests per year scenarios. In aggregate, incremental testing and treatment costs yield US $17.6 million, US $21.6 million, US $17.6 million, and US $14.7 million for one- to four-tests per year.

### Cost-Effectiveness and Sensitivity Analysis

We estimate the costs per DALY averted to be US $464, US $1,190, US $1,762, and US $2,727 for one-, two-, three-, and four-tests per year scenarios, respectively. All but the four-test per year scenario ICER estimates are less than the estimated opportunity-cost based threshold of US $2,255.

From our probabilistic sensitivity analysis, we find our results are robust to a large variety of key input parameters (Figure 4). We find that 97% of modeled scenarios for one-test would be cost-effective, while 96%, 99.9%, and 100% of two-, three-, and four-tests per year scenarios are cost-effective from our sensitivity analysis.

**Figure 4:**
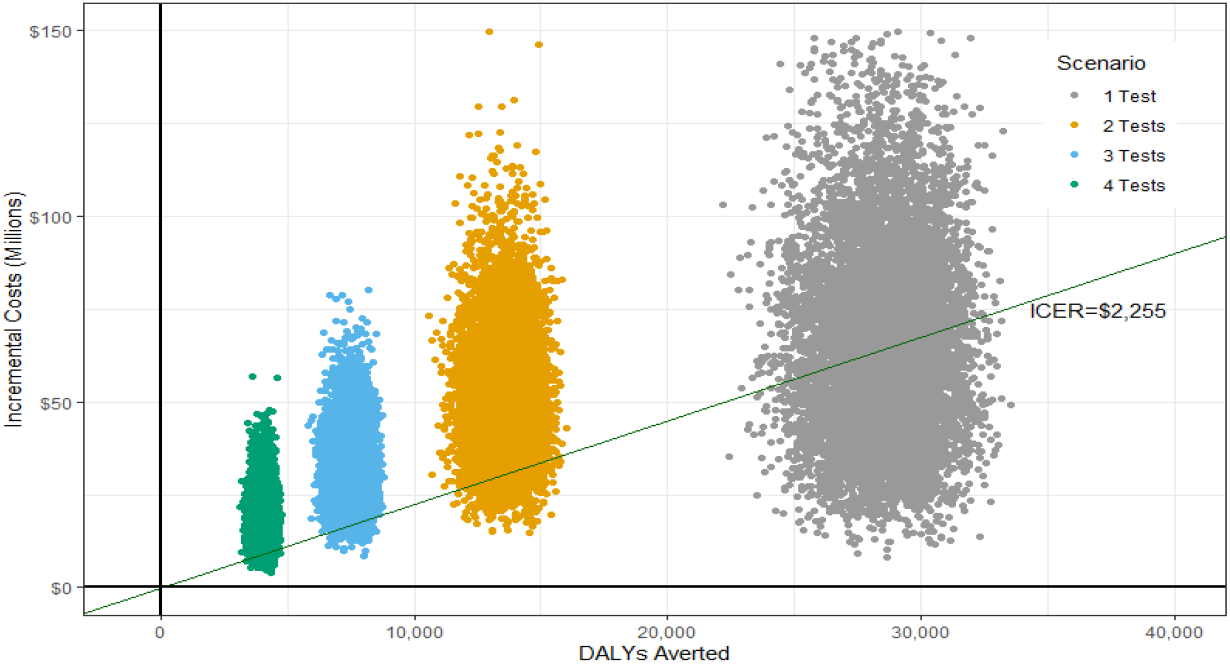
Probabilistic Sensitivity Analysis Results by Test Policy Scenario DALY, disability-adjusted life year; ICER, incremental cost-effectiveness ratio.

We focus on cost of HIV testing and treatment, and health impact of increased testing frequency as parameters of uncertainty and present the associated ICERs by scenario (Figure 5). On a univariate basis, impact could fall by 80% and the one-test scenario would remain cost-effective (all else equal). Conversely, with impact estimates held stable, the ICER for the one-test scenario remain cost-effective even if costs increase by 3 times for the (all else equal). In the two-test scenario, latitude of cost-effective decreases to a 50% decrease in impact or 80% increase in cost. Three-test scenario only remain cost-effective if impact does not drop beyond 20% of our estimate and costs do no increase by more than 20%.

**Figure 5:**
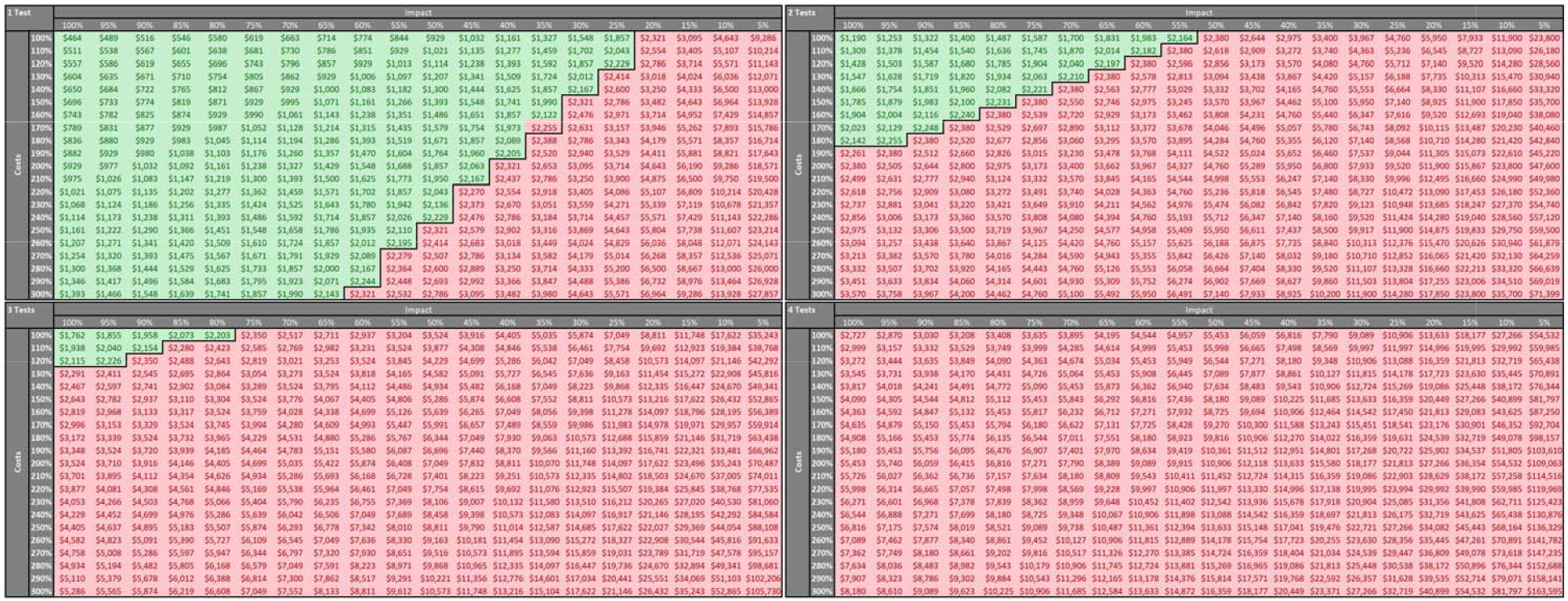
Two Way Sensitivity of ICER per DALY Averted by Impact, Costs, and Test Policy Scenario DALY, disability-adjusted life year; ICER, incremental cost-effectiveness ratio.

## Discussion

We estimated the costs and health impact associated with greater HIV testing frequency among high-risk groups in Viet Nam. We found that scaling up more frequent testing among KP could avert a considerable HIV burden in alignment with expectations surrounding disproportionate risk attributed to KP.^23^ We estimate between 1.6%-10.2% of HIV infections and 0.8%-4.9% of deaths by 2030 in the whole population for HIV testing one- to four-times per year among KP. Testing one- to four-times per year would cost US $464-$2,727 per DALY averted. This health benefit would be achieved by accelerating HIV diagnosis and ART scale-up, as we estimate a policy testing four times per year could reach 95% ART coverage by 2024, rather than 2030 under the status quo testing uptake.

Knowledge of HIV status remains a persistent challenge globally, with only 81% of persons living with HIV estimated to know their status, while only 63% and 75% are estimated to know their status in Asia and Eastern Europe, respectively – regions where HIV incidence is disproportionally comprised of key populations like Viet Nam.^24^ Late presentation (diagnosis with CD4 <200 cells/µL) remains common, leading to higher rates of morbidity and mortality, and more downstream transmission. Those diagnosed late cite feeling healthy, stigma or fear, and inconvenience as reasons to delay testing.^25^ Higher uptake along with increased frequency of testing through differentiated testing modalities could further help close the gap to reaching the UNAIDS targets and national targets.^19^

Recently, community-led HIV self-testing, index testing, and lay provider testing have been found to have high levels of acceptability and uptake and offer further options for individuals in need of HIV testing services.^26–28^ Nevertheless, more frequent HIV testing and faster scale-up of ART alone is unlikely to be sufficient to reach virtual elimination of HIV.^29^ Other modeling efforts have suggested that even universal ART coverage by 6 months following infection can leave notable levels of residual incidence from transmission during the acute infection period.^30^ Therefore, further scaling up HIV prevention services, facilitated through increased HIV testing, will have an impact on both HIV prevention and mortality reduction. Inclusion of additional testing modalities such as index and self-testing could build upon this impact.

Our study has several limitations. First, there is limited quantitative information on the empirical link between HIV testing frequency and ART uptake. However, we made a conservative assumption about this relationship, and ultimately believe this link is plausible, necessary, and strong. Second, our cost assumptions are based on local government data, and reflect current service delivery modalities, efficiencies, and economies of scale and scope. Future costs of ART may continue to go down, as has been well documented over the past 30 years, however some speculate that we are near the cost floor for treatment.^31,32^ In contrast, HIV testing may get more expensive, as bespoke testing modalities which target individuals not served by existing approaches may be more expensive on a per person basis. Alternatively, more frequent testing could benefit from economies of scale and marginal costs could decrease.^33^ Nevertheless, our estimates suggest that more frequent HIV testing would remain cost-effective, even in scenarios where per person testing costs increased by nearly three-fold, implying the normative findings of cost-effectiveness are likely to remain unchanged even if more expensive testing models are needed to increase overall coverage and frequency. Finally, we made assumptions regarding the uptake and utilization of HIV testing services. Preferences on the timing and frequency of HIV testing by population is unknown in the aggregate, and at the individual level could vary tremendously over time. We made conservative assumptions about testing uptake and assume that at least a quarter of the target population may be reluctant to test, irrespective of policy.

Progress towards the first 95 of the UNAIDS targets is essential to achieving global and local goals of reducing the health burden and inequity of HIV and maintaining low HIV incidence. We believe that offering more frequent HIV testing services for high-risk populations could be an instrumental, cost-effective, and affordable policy option. Further research should be focused on strategies to meet the preferences of clients’ testing frequency, timing, and type, ensure these strategies are of high quality with high levels of linkage and retention, as well establish better empirical estimates of the cost-outcome curve associated with more HIV testing.

## Data Availability

All data produced in the present study are available upon reasonable request to the authors

